# Predicting monopolar local field potential power from bipolar recordings in deep brain stimulation

**DOI:** 10.64898/2026.03.13.26348345

**Authors:** Chance Fleeting, Griffin Lamp, Kara A. Johnson, Jackson N. Cagle, Coralie de Hemptinne, Aysegul Gunduz, Joshua K. Wong

## Abstract

**Objectives:** Deep brain stimulation (DBS) is an established therapy for neurological disorders such as Parkinson’s disease (PD). Modern DBS devices can record local field potentials (LFPs) to guide DBS therapy. LFPs from these devices are typically limited to bipolar configurations to suppress common-mode noise and reject artifacts. However, bipolar recordings also attenuate some local physiological signals. Methods that convert bipolar to monopolar power offer more spatially precise estimates of LFPs. Herein, we develop a model to estimate monopolar power from bipolar recordings.

**Materials and Methods:** This retrospective study analyzed 64 leads in 50 patients with PD undergoing STN (11) or GPi (53) DBS implantation. Intraoperatively, LFPs were recorded from all contacts and filtered. Bipolar montages were generated for each combination. Power spectral density (PSD) was calculated from each monopolar and bipolar signal, averaged over canonical frequency bands, and processed as log PSD. A common set of bipolar configurations was selected to minimize the Condition Number (CN), maximizing model stability. Monopolar and bipolar powers were related using robust OLS regression. Observations were randomly partitioned into training and validation sets.

**Results:** Sixty-four leads yielded 640 observations. The configuration with the lowest CN (7.45) was {C_03_, C_12_, C_23_}. The models demonstrated adjusted R^2^s of 0.9015, 0.9055, 0.8853, and 0.8764, and RMSEs (dB) of 3.2663, 3.2801, 3.5815, and 3.7035 when predicting C0, C1, C2, and C3 (N = 500; all p < 0.0001). Weights transferred from the training set to the validation set, as well as STN, GPi, and hemisphere-specific subsets, retained high performance.

**Conclusions:** This study demonstrates that monopolar LFP power can be accurately estimated from bipolar power using a linear regression model with strong generalizability across targets and validation sets. This approach offers a hardware-agnostic solution to spatially disambiguate signals and better inform DBS programming and adaptive stimulation in chronically implanted devices.

## Introduction

Deep brain stimulation (DBS) is an established therapy for movement disorders such as Parkinson’s disease (PD), essential tremor, and dystonia^1–3^. Contemporary DBS devices can record local field potentials (LFPs), facilitating real-time assessment of brain states^4^, which can guide DBS therapy^5,6^.

Currently, LFPs from implanted DBS devices are typically limited to bipolar configurations between two contacts, suppressing common-mode noise. This is particularly useful during active stimulation when large artifacts can obscure physiological activity. However, measuring the potential difference between closely spaced contacts also attenuates physiological signals. Thus, bipolar recordings limit spatial resolution and sensitivity to local common-mode neuronal activity compared to monopolar recordings referenced to distant electrodes. Establishing a method to convert bipolar to monopolar power could yield more spatially precise estimates of LFP activity, thereby more effectively guiding DBS therapy.

In this study, intraoperative LFPs were acquired in patients undergoing DBS implantation for PD. The recordings were acquired in a monopolar configuration by temporarily connecting the DBS leads to an external neuroamplifier. A montage of bipolar LFPs was then constructed from these interoperative recordings to function as a simultaneous signal. The original monopolar LFPs were used as the ground truth to identify the relationship between bipolar and monopolar signals, yielding a method for estimating monopolar power from bipolar recordings, with significant implications for LFP-guided DBS therapy.

## Materials and Methods

This retrospective study analyzed data from 64 leads representing 50 patients with PD undergoing subthalamic nucleus (STN; n=11) or globus pallidus internus (GPi; n=53) DBS implantation at the University of Florida. This study was approved by the University of Florida IRB (IRB202202848).

Following lead placement, 60-second LFP recordings were obtained at 22 kHz (Neuro Omega, Alpha Omega, Israel), capturing activity from all contacts (C_0_, C_1_, C_2_, and C_3_) referenced to a scalp corkscrew electrode. Patients were awake, at rest, and off dopaminergic medications ≥ 12 hours before surgery. All patients were implanted with quadripolar DBS leads (Medtronic Models 3387 or 3389, Minneapolis, MN).

In each patient’s LFPs, a 30-second window free of gross artifacts, such as movement or drop-out artifact, across all contacts was isolated via visual inspection and filtered (5^th^-order Butterworth; band-pass 1-500 Hz) to remove drift and noise. Line noise was notch-filtered at 60 Hz. Physiological artifacts, such as ECG contamination, were not explicitly filtered but did not make a notable visual contribution to the collected signals. To compute bipolar montages (C_01_, C_02_, C_03_, C_12_, C_13_, and C_23_), differences were taken between the indicated monopolar LFP recordings (e.g., the bipolar signal at C_01_ is equivalent to the monopolar signal at C_0_ subtracted from that at C_1_). For both monopolar and bipolar recordings, power spectral density (PSD) was computed using Welch’s method (1-second Hann window, 0.5-second overlap; SciPy welch).

Each bipolar and monopolar power was then averaged over 10 canonical frequency bands: delta (0.5-4Hz), theta (4-8Hz), alpha (8-12Hz), beta (13-30Hz), low beta (13-20Hz), high beta (20-30Hz), gamma (30-80Hz), low gamma (30-55Hz), high gamma (55-80Hz), high frequency oscillations (>80Hz). Values were processed as log PSD, referenced to 1 μV^2^/Hz. For each patient and electrode, the monopolar powers of the 10 frequency bands were paired with the corresponding powers from 3 of 6 possible bipolar configurations (Fig 1A) in a given observation. The specific group of 3 bipolar configurations used across all recordings was selected by maximizing the stability of the linear regression model by minimizing the Condition Number (CN), a metric that uses eigenvalues to assess input-variable multicollinearity and shared information to predict how drastically outputs vary in response to small input perturbations^7^. CN was limited to CN < 10 to avoid multicollinearity^8^. Sets with higher CNs risk inflated and inaccurate significance and goodness-of-fit.

**Figure 1.**
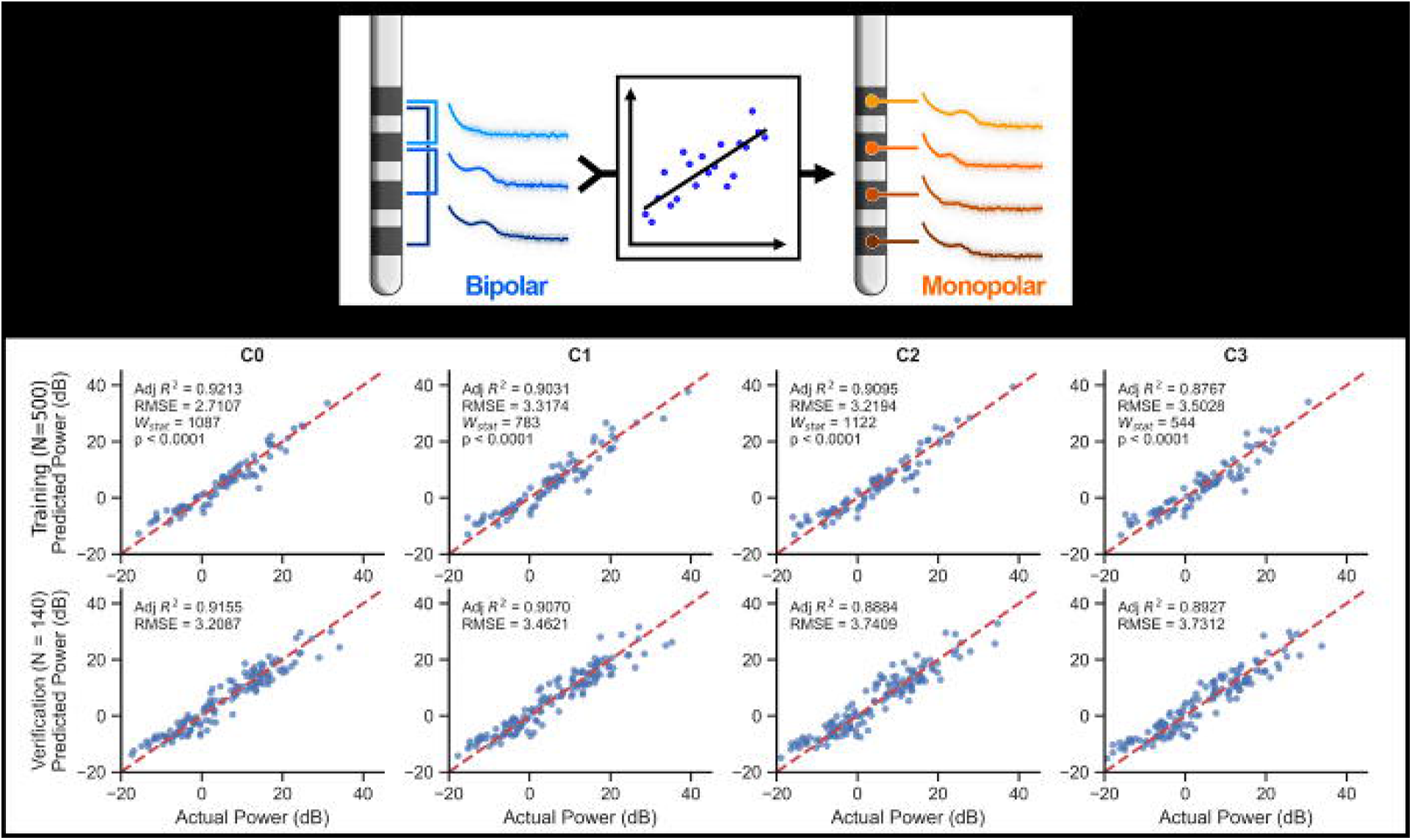
Estimation of bipolar local field potential power to a monopolar power. (A) A cartoon showing that multiple regression models translate multiple bipolar local field potential power readings into a given monopolar power. (B; First Row) Scatter plots comparing actual and predicted values from the training set for each of the four monopolar contacts show a strong correlation, indicating a robust model. (B; Second Row) Scatter plots comparing the actual and predicted values from the validation set show a similarly strong fit, implying that the weight may generalize.

For this analysis, the monopolar powers were the dependent variable, and each of the three corresponding bipolar configurations served as an independent variable. This set size (3) was chosen empirically using CN but is also consistent with the rank expected to span a 4-contact bipolar space. The monopolar and bipolar powers for each observation were related using ordinary least squares (OLS) regression. To account for heteroskedasticity and autocorrelation, variance was estimated using Newey-West (HAC) standard error with a 1-sample lag. Regression was performed using Statsmodels (v0.14.2) in Python (v3.12).

The collected observations were randomly partitioned into 500 for training the regression model, with the remainder used for validation.

## Results

Sixty-four leads (11 STN and 53 GPi) from 50 patients with PD were included, with simultaneous LFP powers for each configuration across all 10 canonical bands, yielding 640 observations. The configuration group associated with the lowest CN (7.45) was {C_03_, C_12_, C_23_}. The configuration that matched the default recording paradigm from the Medtronic Percept system, {C_02_, C_03_, C_13_}, exceeded our multicollinearity threshold (CN = 12.39) and was thus excluded from further analyses. The dataset was randomly partitioned into 500 observations for training/fitting and 140 for validation.

The models demonstrated adjusted R^2^s of 0.9015, 0.9055, 0.8853, and 0.8764 when predicting C_0_, C_1_, C_2_, and C_3_, respectively, using the training data (N = 500; all p < 0.0001 via Wald test; Fig 1B). The regression model coefficients are shown in Table 1. The root-mean-square errors (RMSE; dB) of the regression models were 3.2663, 3.2801, 3.5815, and 3.7035, respectively.

**Table 1.**
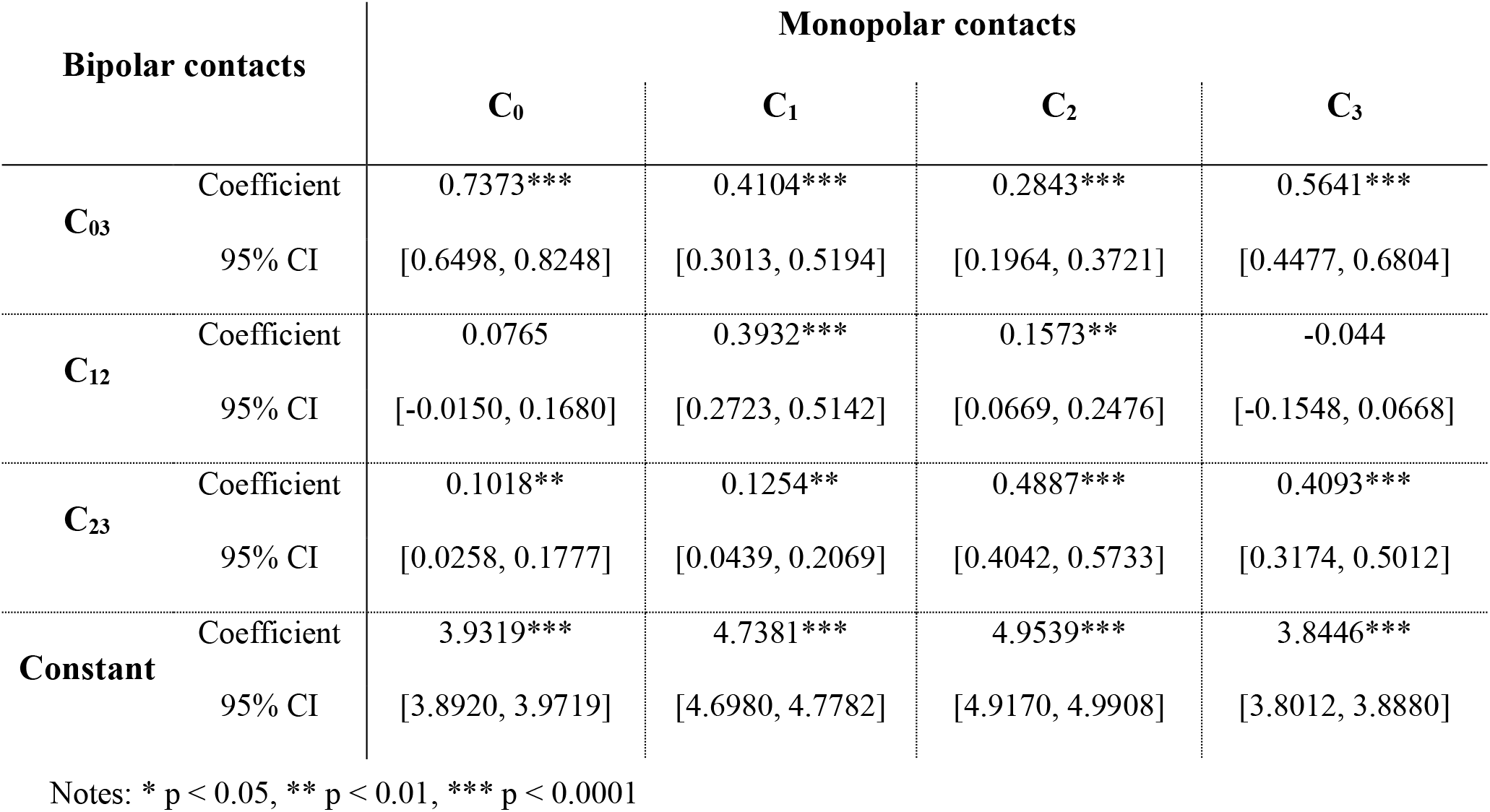
Linear weights to estimate monopolar LFP power from bipolar LFP power.

The weights obtained from this model were transferred and applied to the validation set without adjustment. This resulted in naïve adjusted R^2^s of 0.9155, 0.9070, 0.8884, and 0.8927 and RMSEs of 3.2087, 3.4621, 3.7409, and 3.7312, respectively (N = 140; Fig 1B).

After dividing the validation set into GPi-only and STN-only groups, as well as left- and right-hemispheres, the same transfer-validation methods were applied. Using GPi-only (N = 116), the adjusted R^2^s were 0.8990, 0.9080, 0.8842, and 0.8805 with RMSEs of 3.3389, 3.2175, 3.5712, and 3.6641, respectively. Similarly, STN-only (N = 24) adjusted R^2^s were 0.9036, 0.8115, 0.8705, and 0.8002 with RMSEs of 2.8230, 4.230, 3.728, and 4.208, respectively. The left-(N = 68) and right-hemisphere (N = 72) validation groups showed similar results, as shown in Figure S3. All p-values were < 0.0001 via permutation testing. Performance was comparable across both sub-cohorts and when the sub-cohorts were combined.

A companion script containing a pretrained linear model using the weights from this article has been made available^9^. For examples of clinical applications, please see the companion script.

## Discussion

This study presents the first analysis of a dataset comprising simultaneous monopolar and bipolar LFP recordings acquired from DBS leads to create an empirical method to convert from bipolar to monopolar power. We derived multiple regression models for each contact, demonstrating high goodness-of-fit in predicting monopolar power from bipolar recordings. These models provide a novel approach to localize LFP activity, enabling spatial differentiation of anode and cathode contributions in bipolar recordings. The implications of these models are significant for LFP-guided DBS programming, as monopolar recordings provide more directly actionable information during routine DBS programming. As an inverse problem, current interpretations of bipolar recordings rely on mathematical and theoretical frameworks^10,11^. This study, however, leverages intraoperative recordings and monopolar data to detail the relationship between bipolar power and monopolar contact power. This complements the theoretical work to estimate the surrogate, such as pseudomonopolar LFPs^12,13^, by filling in gaps provided by a pure geometric approach that otherwise prohibit its use within the near-field of quadripolar leads.

When analyzing the validation set with pretrained coefficients, the system maintained a strong fit, suggesting the coefficients generalize well. However, the training-validation partition was randomized without controlling or matching patient parameters. This method was chosen to maximize subject representation when generating weights, since, from a signal-processing standpoint, each observation is expected to be mathematically independent. Additionally, since each observation was indexed by patient and frequency band, observations in the training and validation sets may share related sources. Repeated analysis and validation with strict but randomized partitioning of patients yielded similar results (Figure S4). It should be noted that STN vs. GPi representation in the training and validation set was attempted but was unable to be achieved perfectly due to the number of patients represented in this study. Further, this patient-wise partition means that the weights captured by this alternative method represent only 80% of the patient cohort.

Future work is needed to provide external validation of data collected from patients outside this cohort, including *in situ* simultaneous direct measurement of mono-versus bipolar LFPs, to confirm transferability to general application and generalizability across larger populations. This would help identify any systematic differences between reconstructed and directly measured signals that we would otherwise be unable to detect. Additional future work is required to explore more sophisticated, nonlinear iterations of this model. The increased statistical power and flexibility of these methods would enable expansion into narrow-band and continuous-spectrum analyses, further improving the clinical versatility of this model. Following this, it is possible that reconstructed monopolar LFPs may offer additional benefits we have not yet considered, such as coupling the artifact-rejection power of bipolar recordings with the targeting-relevant specificity of single-contact measurements. Investigating avenues such as these will require substantially higher statistical power, representation, and access to *in situ* simultaneous signal acquisition.

Validating the model on individual targets (STN-only and GPi-only), we observe similarly strong predictive values. While the R^2^ and RMSE are comparable to the full cohort, smaller sample sizes weaken the law of large numbers and increase reliance on normality assumptions, which are likely violated. The comparable performance when verified on the full cohort versus individual subcohorts suggests physiological differences between STN and GPi do not significantly affect the predictability of monopolar LFP power. This is important as other electrophysiological characteristics, such as evoked potentials, vary greatly between STN and GPi^14,15^. Given the strong performance using the full cohort, a single unified model appears viable, which is preferable for generalized analyses.

Previous research established strong correlations between brain electrophysiology and PD motor symptoms, leading to the development of electrophysiology-guided DBS programming^16–20^. Many studies used intraoperative recording systems capable of monopolar LFP acquisition, but translating these findings to chronic DBS devices is challenging because of the spatial ambiguity introduced by the inability to differentiate cathode and anode contributions in bipolar recordings.

While recent hardware advancements, such as the electrode identifier (EI) in Medtronic Percept, allow physicians and researchers to read monopolar LFPs *in situ*, they generally require specific hardware requirements, such as bilateral implantation, to provide a common anode via the contralateral lead. With this setup, Percept directly measures monopolar signals, which carry the same risks of leakage and interference from erroneously propagated signals from the common reference, similar to ECG leakage in the traditional monopolar case. Our linear model offers a translatable solution for chronic DBS devices that would otherwise only enable bipolar power measurements, facilitating broader, more precise adaptive DBS protocols and more robust study designs. This bypasses concerns regarding the precision of bipolar LFP-based localization of neural biomarkers^21,22^.

In practice, this model can be applied directly to postoperative sensing recordings by exporting bipolar PSD values from the device interface (e.g., BrainSense survey), extracting the required bipolar channels {C_03_, C_12_, C_23_}, and then inputting these values into the provided companion script. The script then estimates the monopolar powers for each individual contact. No additional preprocessing is required, and this should fit within most users’ standard PSD pipelines. Theoretically, other bipolar configurations, such as the default {C_02_, C_03_, C_13_}, can be converted into the set requested prior to power calculation via subtraction of corresponding voltages (re-referencing; e.g. V_23_ = V_03_ - V_02_); however, this propagates uncertainty and assumes that the user has access to raw voltages.

## Conclusions

Using the weights found herein, monopolar LFP power can be estimated from bipolar power to differentiate the origin of the electrophysiologic biomarker and identify the most proximal contact to given neurological activity. These findings can directly inform clinicians’ and researchers’ selection of programming configuration for a given neuromodulation scenario.

## Data Availability

All data produced in the present study are available upon reasonable request to the authors

https://chancefleeting.github.io/Research/Bipolar2Monopolar/Fleeting_LFP_MonoPowerEstimate.html

## Acknowledgements

All authors have no relevant conflicts of interest with the present work. AG received Medtronic devices under an NIH BRAIN Public-Private Partnership used for intraoperative recordings in the study. JKW was supported by NIH KL2TR001429. KAJ was supported by NIH K99NS137249. CDH received research support from the Parkinson’s Foundation. CF, GL, AG, and JNC report no financial disclosures.

## Author Roles

CF: conceptualization, analysis, writing, editing of final version of the manuscript; GL: conceptualization, analysis, editing of final version of the manuscript; KAJ: conceptualization, analysis, editing of final version of the manuscript; JNC: conceptualization, analysis, editing of final version of the manuscript; CDH: conceptualization, editing of final version of the manuscript; AG: conceptualization, editing of final version of the manuscript; JKW: conceptualization, design, execution, analysis, writing, editing of final version of the manuscript

## Data availability

The data supporting the findings of this study are available from the corresponding author upon reasonable request.

## Supplemental Legend

**Figure S1. KDEs of power distributions for each LFPs.** Since the graphs are not normally distributed, a large sample size is required to attain a reliable model.

**Figure S2. CN distribution for all contact groupings of 3 or more bipolar configurations from the six standards.** A CN of 10 was the threshold for moderate multicollinearity, while 30 or higher indicated severe multicollinearity.

**Figure S3. Scatterplots displaying the fit transferred to validation sets,** including GPI-only, STN-only, left-, and right-hemisphere subgroups.

**Figure S4. Estimation of bipolar local field potential power to a monopolar power** fit using a strict patient-wise division between the training and validation set.

