## Supplemental Figures for "Predicting monopolar local field potential power from bipolar recordings in deep brain stimulation"

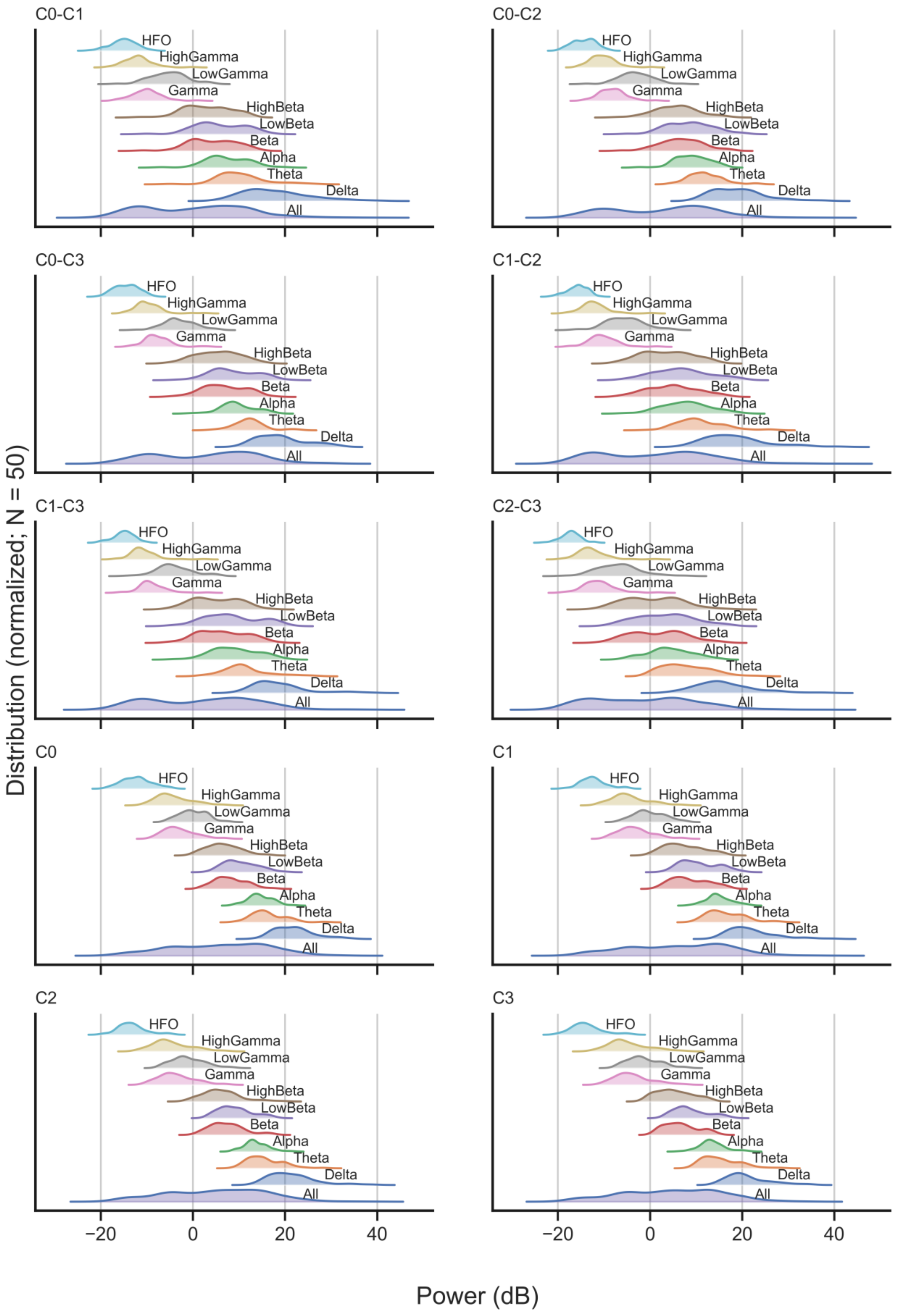


**Figure S1. KDEs of power distributions for each LFPs.** Since the graphs are not normally distributed, a large sample size is required to attain a reliable model.

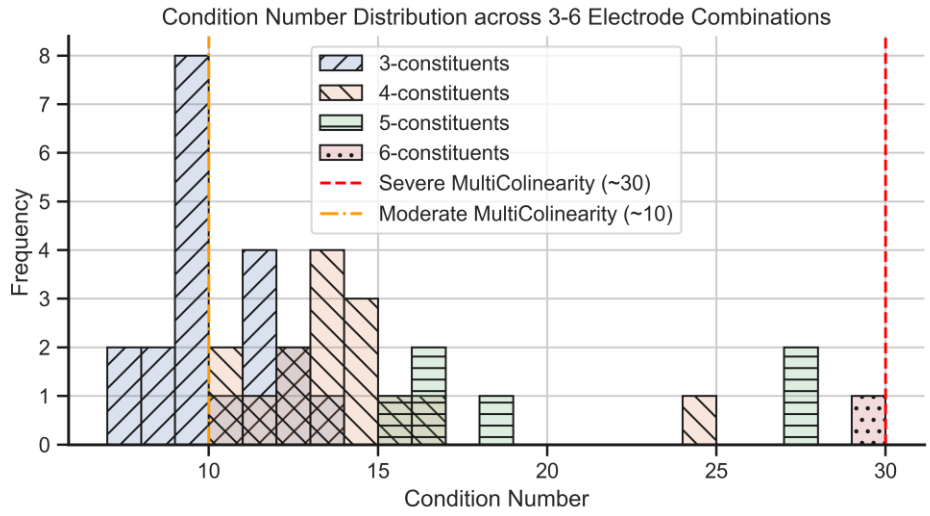


**Figure S2. CN distribution for all contact groupings of 3 or more bipolar configurations from the six standards.** A CN of 10 was the threshold for moderate multicollinearity, while 30 or higher indicated severe multicollinearity.

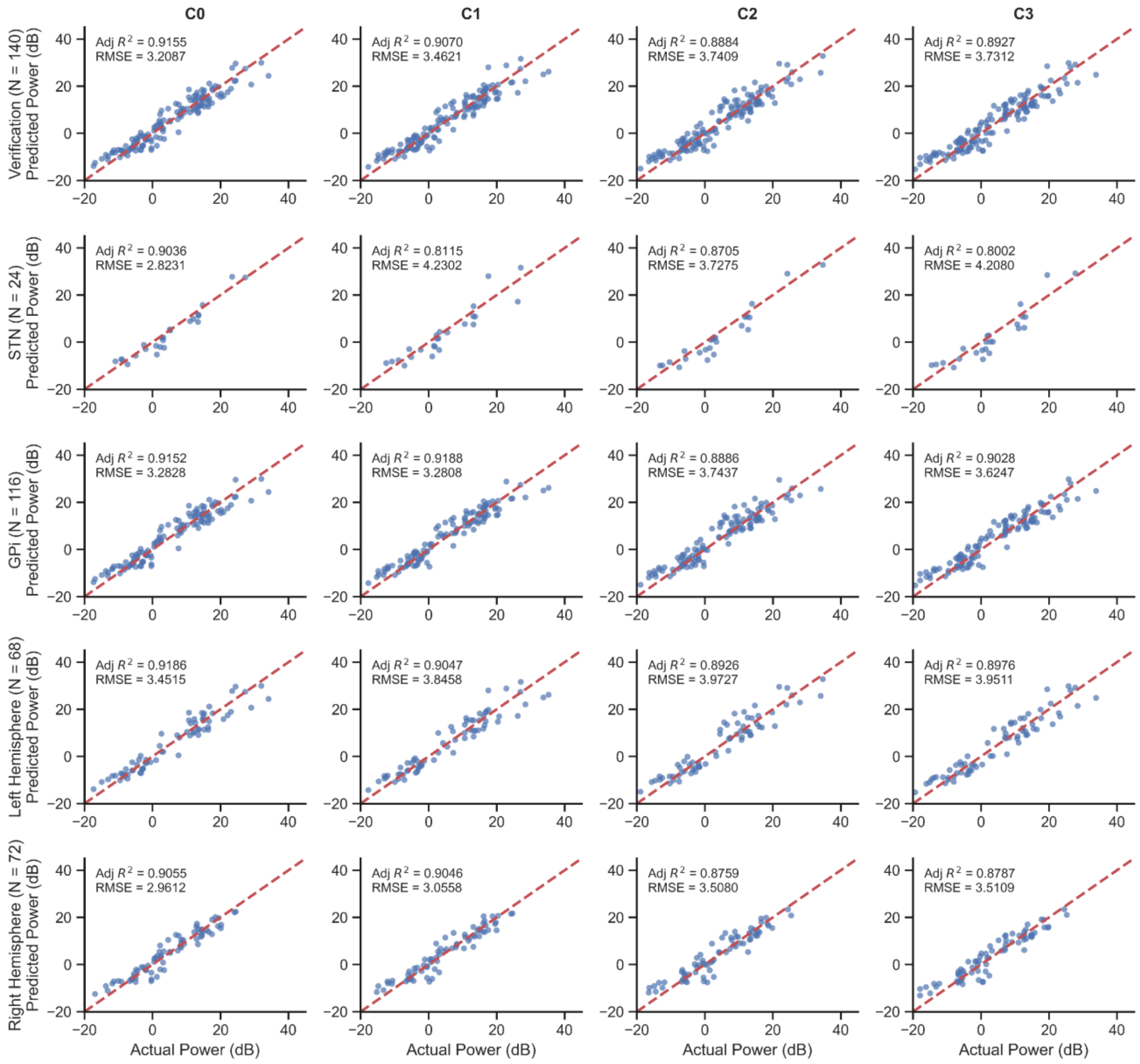


**Figure S3. Scatterplots displaying the fit transferred to validation sets,** including GPI-only, STN-only, left-, and right-hemisphere subgroups.

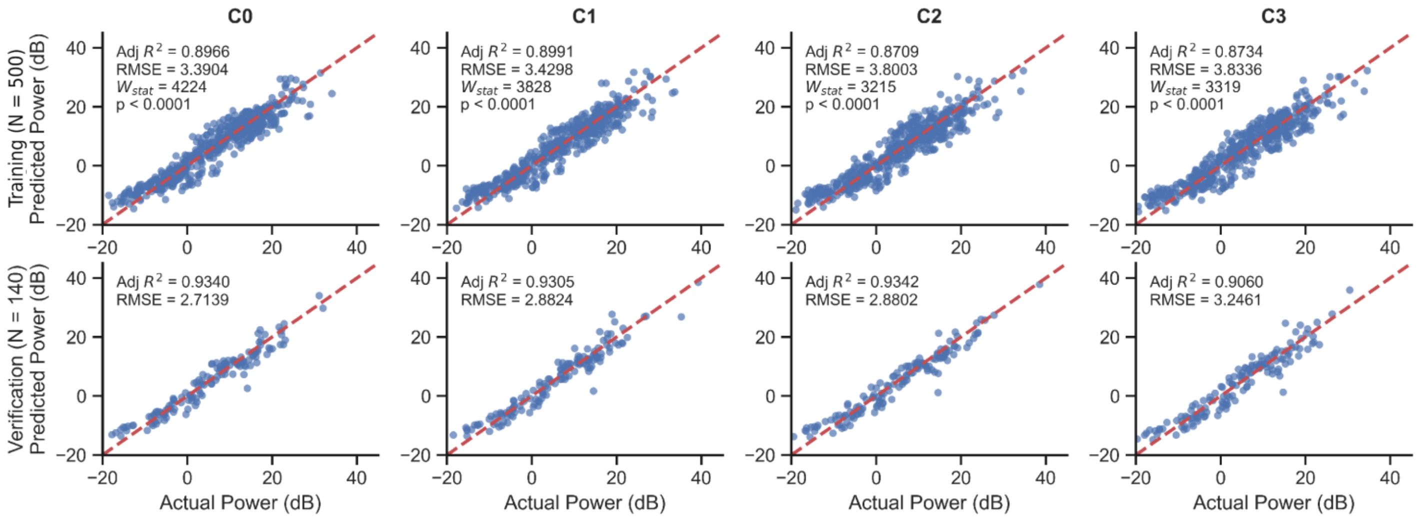


**Figure S4. Estimation of bipolar local field potential power to a monopolar power** fit using a strict patient-wise division between the training and validation set.
